# The EEG–Respiratory Gap: a deep learning biomarker associated with long-term mortality in obstructive sleep apnea

**DOI:** 10.64898/2026.11.22.26368628

**Authors:** Hyun Kyung Lee, Keondo Park, You Rim Choi, Da-eun Lee, Hyung-Sin Kim, Hyun-Woo Shin

**Affiliations:** Obstructive Upper Airway Research (OUaR) Laboratory, Department of Pharmacology, Seoul National University College of Medicine, Seoul, Republic of Korea; Graduate School of Data Science, Seoul National University, Seoul, Republic of Korea; Sensory Organ Research Institute, Seoul National University College of Medicine, Seoul, Republic of Korea; OUaR LaB, Inc, Seoul, Republic of Korea; Department of Otorhinolaryngology-Head and Neck Surgery, Seoul National University Hospital, Seoul, Republic of Korea; Ischemic Hypoxic Disease Institute, Seoul National University Medical Research Center, Seoul, Republic of Korea; Cancer Research Institute, Seoul National University College of Medicine, Seoul, Republic of Korea

## Abstract

The apnea-hypopnea index (AHI) summarizes respiratory events by frequency, but was not designed to reflect how strongly an individual’s brain responds to them. We developed a deep-learning model that estimates AHI from a single EEG channel (AHI_brain_), developed using 22,044 polysomnography recordings and validated in an internal test set (*r* = 0.88) and three external test sets (*r* = 0.50–0.62). From AHI_brain_, we derived two complementary metrics that characterize the EEG–Respiratory Gap: ΔAHI_brain_, reflecting whether a patient’s AHI_brain_ is higher or lower than expected for their measured AHI, and AHI_brain,event-free_, estimated from epochs without scored respiratory events. Both were associated with sleep macro-, micro-, and epoch-level EEG features, suggesting that this discrepancy is not solely attributable to model error. Among participants free of cardiovascular disease, both metrics were associated with all-cause and non-CVD mortality after adjustment for AHI and clinical risk factors. After accounting for the same covariates, AHI_brain,event-free_ was also associated with CVD mortality in the highest-versus-lowest quartile comparison. These findings support the EEG–Respiratory Gap as an interpretable EEG-derived measure associated with prognostic information beyond AHI.

## Introduction

Obstructive sleep apnea (OSA) is estimated to affect nearly one billion people worldwide^1^ and is associated with long-term mortality^2^, yet the optimal metric for assessing its severity remains unclear^3^. The apnea-hypopnea index (AHI) remains the most widely used severity metric and summarizes OSA primarily by the frequency of scored apnea and hypopnea events. However, individuals with similar AHI can differ in the physiological consequences associated with those events^3–5^. This heterogeneity has motivated complementary measures that characterize dimensions of OSA not represented by event frequency alone. For example, ventilatory and hypoxic burden measures incorporate information from airflow reduction or event-related hypoxemia and have shown associations with cardiovascular and mortality outcomes beyond those observed with AHI^6^. These measures provide a richer characterization of respiratory and oxygenation burden, but remain primarily derived from respiratory or oximetric signals^7–9^.

Respiratory disturbances may also impose cumulative physiological burden on end organs, particularly the brain. OSA has been associated with accelerated cognitive decline and an increased risk of mild cognitive impairment and dementia^10–12^. Neurocognitive vulnerability also varies across individuals and is influenced by traits such as the respiratory arousal threshold, which reflects the propensity to arouse in response to a respiratory event, rather than by event frequency alone^13,14^. Consistent with the potential importance of arousal-related physiology, arousal burden, defined as the cumulative duration of scored arousals normalized to total sleep time, has been associated with long-term all-cause and cardiovascular mortality in MrOS and SHHS, cohorts also analyzed in the present study^15^. Respiratory drive and arousal responses to airway obstruction are generated and modulated by brainstem and cortical circuits^16,17^. EEG may therefore provide a means to characterize neural responses associated with respiratory stress, complementing conventional metrics derived from respiratory signals.

Several studies have shown that AHI can be estimated from EEG alone. Alongside broader deep-learning approaches to AHI prediction^18,19^, several studies have used EEG to detect respiratory events or estimate AHI. Kokkalas et al.^20^ detected individual apnea events with a convolutional-recurrent network and reported strong agreement between estimated and polysomnography-derived AHI (*r≈* 0.88). Cheng et al.^21^ jointly modeled sleep staging and OSA-event detection, while Wan et al.^22^ combined a CNN, Transformer, and CRF to link sleep-stage sequences with OSA-severity regression. Barnes et al.^23^ used an explainable CNN for single-channel EEG apnea detection and Mahmud et al.^24^ detected apnea from sub-frame EEG feature variation. However, in these studies, estimation accuracy was the primary goal, and the discrepancy between the EEG-derived estimate and the measured AHI was treated as residual error rather than as information. The present study takes the comparable internal agreement (*r* = 0.88) as a starting point rather than an endpoint and asks what information this discrepancy itself may contain.

Treating a predicted-observed discrepancy as an individual vulnerability marker, rather than noise, is an established strategy across modalities and target variables. A sleep-EEG-derived brain-age index – predicted minus chronological age – independently predicted reduced life expectancy in SHHS^25^. A multimodal-PSG extension similarly linked age-estimate error to all-cause and cardiovascular mortality^26^. The same logic extends beyond sleep and age: ECG-based BMI-estimate error independently predicted future cardiometabolic disease^27^, while ECG-based age-estimate error independently predicted mortality^28^. Mortality provides an objective, clinically consequential endpoint with long-term follow-up available in large, well-characterized cohorts, making it a natural first test of the prognostic relevance of such a discrepancy. We apply this logic to OSA, defining the EEG–Respiratory Gap as the discrepancy between EEG-estimated (AHI_brain_) and measured (AHI_GT_) AHI. This construct is characterized using two complementary metrics: ΔAHI_brain_, which quantifies the AHI-adjusted discrepancy between AHI_brain_ and AHI_GT_, and AHI_brain,event-free_, which captures EEG-derived AHI information remaining in epochs without scored respiratory events.

Recent sleep foundation models have demonstrated that EEG and multimodal PSG contain clinically relevant information beyond conventional sleep summaries^29–33^. SleepFM, for example, evaluates AHI-based severity classification as a downstream benchmark and separately uses learned representations for disease-risk prediction^29^. However, it does not model AHI as a continuous prediction target or examine whether deviation from conventionally scored AHI is itself informative. Our objective is therefore distinct from broad representation learning or direct risk prediction. By making AHI the explicit prediction target and treating its discrepancy from conventionally scored AHI as the object of study, we ask whether EEG captures a component of respiratory burden not represented by event frequency alone. We test this AHI-referenced construct against established EEG physiology and long-term mortality, providing an interpretable link between conventional OSA severity and latent information encoded in sleep EEG.

This study therefore (1) develops and externally validates a model that estimates AHI from EEG alone, (2) tests whether the resulting EEG–Respiratory Gap reflects sleep-EEG physiology beyond residual model error, and (3) evaluates whether these two complementary metrics are associated with long-term mortality after adjustment for established risk factors.

## Results

### Dataset and baseline model

This study draws on 22,044 polysomnography (PSG) recordings from a multicohort collection comprising five datasets: the diagnostic-night subset of the Korea Image based Sleep Study dataset (KISS_diag_), the Multi-Ethnic Study of Atherosclerosis (MESA), the Osteoporotic Fractures in Men Study (MrOS), the Sleep Heart Health Study (SHHS), and the CPAP titration-night subset of Korea Image based Sleep Study (KISS_cpap_) (Fig. 1a). Records with total sleep time (TST) *<*180 minutes, missing AHI, or missing basic demographic information (age, sex, or BMI) were excluded before partitioning. The Training, Validation, and Internal Test partitions were drawn from pooled KISS_diag_ + MESA + MrOS data and stratified by dataset and AHI severity category (Supplementary Fig. 1a,b). SHHS visits 1 and 2 were held out entirely for external validation. KISS_cpap_ was also held out from model development but was treated as a titration-specific stress-test cohort rather than a conventional diagnostic external-validation cohort, because airway pressure is adjusted during CPAP titration to suppress respiratory events (Table 1, Supplementary Table 1). Repeat visits from the same individual (e.g., the same subject serial number recorded at both MrOS1 and MrOS2) were assigned to a single partition, ensuring that no participant contributed data to both model development and test sets.

**Table 1.** Baseline characteristics of the study population by data split.

| Variable | Train | Valid. | Test | Ext. Test<sup>†</sup> |
|---|---|---|---|---|
| Subject *N* | 8,777 | 1,091 | 1,101 | 7,534 |
| PSG *N* | 9,552 | 1,189 | 1,181 | 10,122 |
| Age, yr | 57.8±19.0 | 58.4±18.8 | 57.0±19.5 | 61.3±13.2 |
| Male, *n*(%) | 7,918(82.9) | 967(81.3) | 985(83.4) | 5,586(55.2) |
| BMI, kg/m<sup>2</sup> | 26.5±4.2 | 26.5±4.4 | 26.3±4.3 | 28.0±5.0 |
| AHI, /hr | 31.0±25.4 | 31.2±24.9 | 31.7±26.0 | 16.4±15.6 |
| TST, min | 325.5±70.2 | 326.9±68.9 | 325.8±71.8 | 350.7±69.7 |
| *OSA severity* |  |  |  |  |
| Normal (<5) | 1,104(11.6) | 137(11.5) | 140(11.9) | 2,297(22.7) |
| Mild (5–15) | 2,100(22.0) | 252(21.2) | 248(21.0) | 3,786(37.4) |
| Mod. (15–30) | 2,353(24.6) | 296(24.9) | 289(24.5) | 2,508(24.8) |
| Severe (≥30) | 3,995(41.8) | 504(42.4) | 504(42.7) | 1,531(15.1) |

Subject *N*, number of unique individuals; PSG *N*, number of polysomnographic studies. <sup>†</sup>External Test comprises the SHHS and KISS<sub>cpap</sub> cohorts.

**Figure 1.**
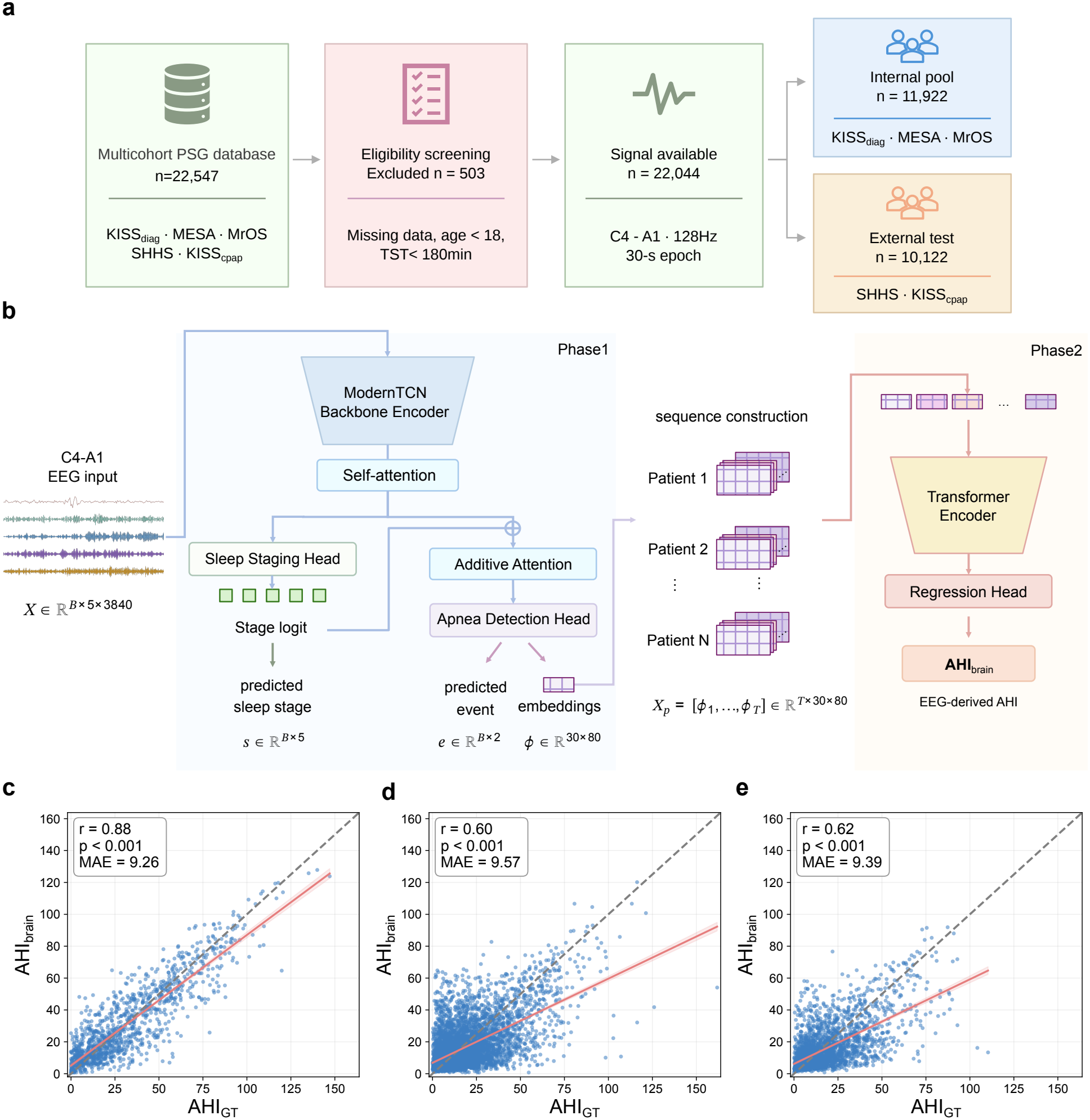
Overview of the two-phase framework for EEG-derived AHI estimation. **a**, Data-flow diagram from the multicohort PSG database through eligibility screening to the signal-available analysis set, split into an Internal pool (KISS_diag_, MESA, MrOS) and an External Test set (SHHS, KISS_cpap_). **b**, brAHInet architecture: Phase 1 performs epoch-level sleep-stage classification and respiratory-event detection from EEG using a ModernTCN backbone and self-attention, with the sleep-stage logits informing respiratory-event detection via additive attention; Phase 2 estimates patient-level AHI_brain_ from the sequence of Phase 1 epoch-level event embeddings using a Transformer encoder. **c**, AHI_GT_ vs. predicted AHI_brain_ in Internal Test. **d**, AHI_GT_ vs. predicted AHI_brain_ in SHHS1. **e**, AHI_GT_ vs. predicted AHI_brain_ in SHHS2. In **c–e**, dashed line, identity; red line and shading, fitted regression with 95% CI.

Across all datasets, a single central EEG derivation (C4-A1) was segmented into 30-second epochs and decomposed into five frequency bands to form the model input. We developed brAHInet (Fig. 1b), a two-phase deep learning framework that maps epoch-level EEG representations to a patient-level EEG-derived AHI. Phase 1 is a multi-task network built on the published ModernTCN architecture^34^, a time-series model that uses large-kernel temporal convolutions to capture temporal patterns across multiple scales. We selected this backbone to model the complex temporal structure of EEG signals. It was trained on the pooled KISS_diag_ + MESA + MrOS Training partition to jointly predict respiratory events and five-class sleep stages for each epoch. For respiratory-event labeling, an epoch was considered positive if it contained at least 10 seconds of apnea or hypopnea. The predicted sleep-stage probabilities are then combined with the epoch-level EEG embeddings through an additive attention module to inform respiratory-event classification. Phase 2 is a Transformer-based regression model that takes the temporally ordered sequence of epoch-level event embeddings from Phase 1 and outputs AHI_brain_, the EEG-derived AHI for each PSG recordings. For comparison, we trained and evaluated the previously published convolutional model (EEG-CLNet^21^) on the same data splits as a baseline for the two epoch-level tasks: respiratory-event detection and sleep-stage classification.

### Model performance

At the epoch level, Phase 1 recovered both respiratory-event and sleep stage information from EEG alone. In Internal Test, macro-F1 was 70.2% for respiratory-event detection and 74.4% for sleep stage classification (Table 2, Supplementary Fig. 3a,b). Across the external validation cohorts, sleep stage classification consistently outperformed the EEG-CLNet baseline, while respiratory-event detection was broadly comparable to the baseline despite some decline from Internal Test. These results indicate that EEG contains substantial information about respiratory-event occurrence that generalizes across independent cohorts.

**Table 2.** Performance of brAHInet and the EEG-CLNet baseline across internal and external test datasets.

| Dataset | Model | Phase 1 |  |  |  |  |  |  |  |  |  |  | Phase 2 |  |  |
|  |  | Respiratory Event Detection |  |  |  | Sleep Staging |  |  |  |  |  |  | AHI Regression |  |  |
|  |  | Acc. | Sens. | Spec. | MF1 | Acc. | F1-W | F1-N1 | F1-N2 | F1-N3 | F1-REM | MF1 | *r* | *P* | MAE |
|---|---|---|---|---|---|---|---|---|---|---|---|---|---|---|---|
| Internal Test | **brAHInet** | 75.49 | **70.37** | 77.07 | **70.15** | **78.32** | **87.40** | **51.69** | **81.13** | **74.18** | **77.67** | **74.41** | 0.88 | <0.001 | 9.26 |
|  | EEG-CLNet | **75.70** | 67.78 | **78.14** | 69.95 | 77.46 | 86.50 | 49.14 | 80.88 | 73.33 | 74.96 | 72.96 | – | – | – |
| SHHS1 | **brAHInet** | 75.77 | **56.45** | 78.66 | 61.37 | **78.25** | **84.41** | **18.85** | **82.57** | **72.90** | **73.29** | **66.40** | 0.60 | <0.001 | 9.57 |
|  | EEG-CLNet | **79.56** | 52.13 | **83.67** | **63.81** | 76.52 | 82.00 | 14.93 | 81.52 | 71.27 | 71.05 | 64.15 | – | – | – |
| SHHS2 | **brAHInet** | 77.92 | 51.73 | 81.83 | 62.20 | **76.56** | **83.92** | **21.39** | **81.58** | **71.02** | **68.00** | **65.18** | 0.62 | <0.001 | 9.39 |
|  | EEG-CLNet | **79.39** | **52.19** | **83.45** | **63.62** | 74.71 | 80.72 | 18.10 | 80.50 | 69.18 | 67.12 | 63.12 | – | – | – |
| KISS<sub>cpap</sub> <sup>†</sup> | **brAHInet** | 61.48 | **78.84** | 60.09 | 48.77 | **76.72** | **81.93** | **51.38** | **78.14** | **79.43** | **81.77** | **74.53** | 0.50 | <0.001 | 16.09 |
|  | EEG-CLNet | **63.01** | 75.86 | **61.98** | **49.42** | 74.66 | 77.45 | 48.68 | 76.73 | 76.26 | 80.05 | 71.83 | – | – | – |

Bold indicates the higher value between brAHInet and EEG-CLNet within each dataset. <sup>†</sup>KISS<sub>cpap</sub> is a CPAP titration-night cohort.

This epoch-level information translated into patient-level estimates of respiratory-event frequency. AHI_brain_ was strongly correlated with AHI_GT_ in Internal Test (*r* = 0.88, *p <* 0.001, MAE = 9.26 events/hour; Fig. 1c). Significant correlations were also observed in the two external diagnostic cohorts, SHHS1 (*r* = 0.60, *p <* 0.001, MAE = 9.57 events/hour) and SHHS2 (*r* = 0.62, *p <* 0.001, MAE = 9.39 events/hour; Fig. 1d,e). SHHS1 and SHHS2 had narrower and lower AHI_GT_ distributions than Internal Test (Supplementary Table 1), consistent with restriction of range. Although correlations were lower in these cohorts, estimation errors remained similar across the three diagnostic cohorts.

KISS_cpap_ was evaluated separately as a CPAP titration-night cohort rather than as a conventional diagnostic external-validation cohort. Agreement was weaker and MAE was higher in this cohort, accompanied by systematic upward bias in AHI_brain_ (Table 2). Together, these results are consistent with prior EEG-based studies showing that respiratory-event frequency is substantially represented in the EEG.

### Two EEG-derived metrics of the respiratory gap

A notable observation in KISS_cpap_ suggested that this EEG representation may extend beyond scored events themselves. KISS_cpap_ consists of CPAP titration-night polysomnograms, in which increasing airway pressure suppresses respiratory events. If AHI_brain_ reflected only these events, its estimates would be expected to decrease accordingly. Instead, AHI_brain_ remained correlated with AHI_GT_ (*r* = 0.50, *p <* 0.001; Fig. 2b) and showed systematic overestimation. The median difference (AHI_brain_-AHI_GT_) was +11.81 events/hour in KISS_cpap_, compared with only +0.33 events/hour in Internal Test, and the discrepancy increased with AHI_GT_ (proportional-bias slope +0.496, *p <* 0.001). Notably, 23.0% of participants with AHI_GT_ *<* 5 events/hour had AHI_brain_ *≥*15 events/hour. In contrast, SHHS1 and SHHS2 showed only small biases ( *−*1.64 and *−*2.40 events/hour; Supplementary Table 2). This discrepancy suggested that EEG may retain respiratory-related information beyond the occurrence of discrete scored events. Motivated by this observation, we defined two complementary metrics from the trained Phase 2 model to characterize this EEG–respiratory discrepancy (Fig. 2a). ΔAHI_brain_ represents the discrepancy between EEG-derived and conventionally measured AHI. A positive ΔAHI_brain_ indicates that AHI_brain_ is higher than expected for a patient’s AHI_GT_, whereas a negative value indicates that it is lower than expected. Rather than using the simple difference between AHI_brain_ and AHI_GT_, we quantified this discrepancy as the residual from regressing AHI_brain_ on AHI_GT_, thereby removing the linear dependence of the discrepancy on measured AHI. By construction, ΔAHI_brain_ is therefore uncorrelated with AHI_GT_ (*r≈* 0.00 in Internal Test; Supplementary Fig. 4a).

**Figure 2.**
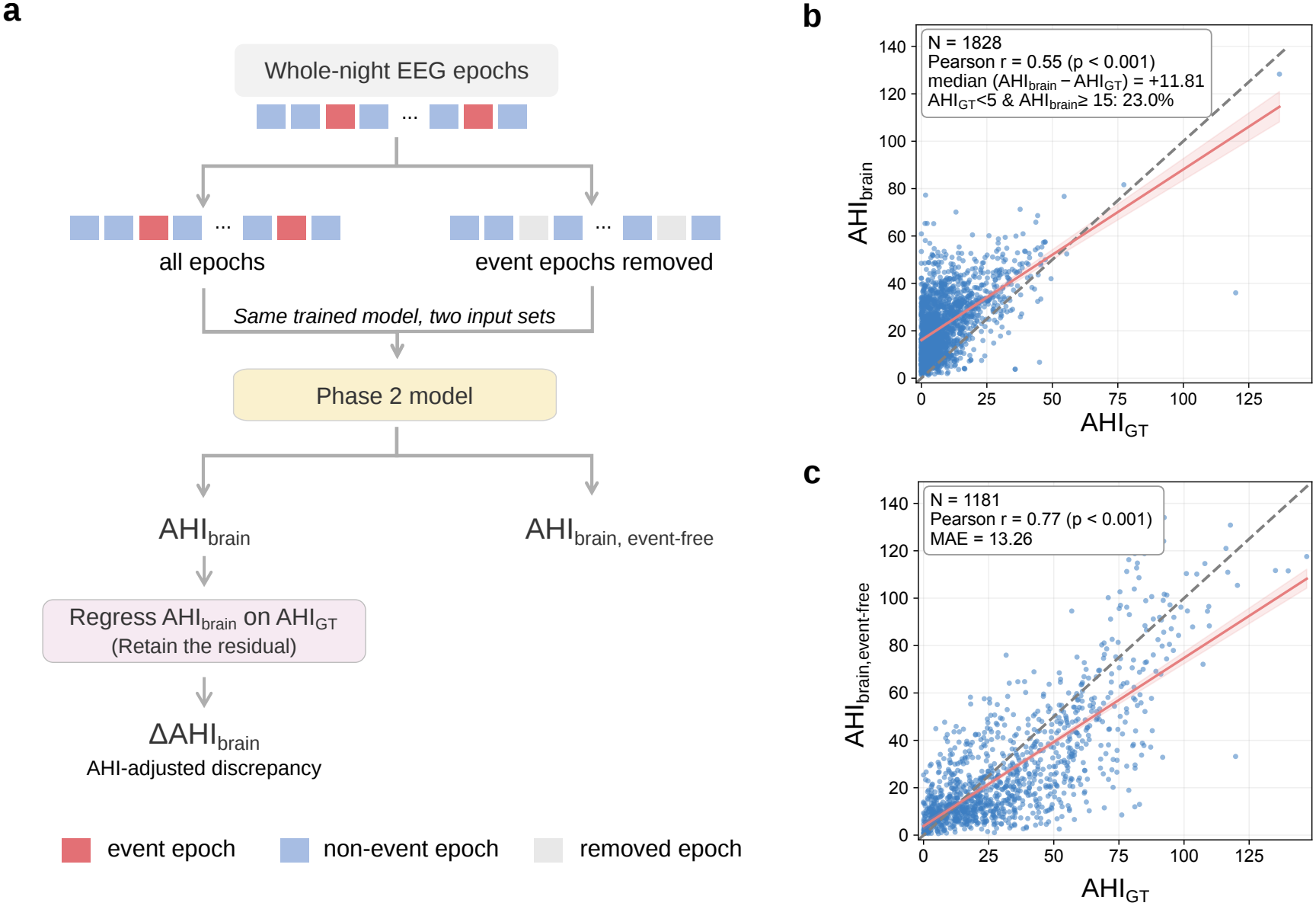
Two EEG-derived metrics that capture complementary components of EEG-derived respiratory information: ΔAHI_brain_ and AHI_brain,event-free_. **a**, Schematic of the two derivation pathways: the same trained Phase 2 model is applied to all epochs (yielding AHI_brain_, regressed on AHI_GT_ to give the residual ΔAHI_brain_) and to the same epochs with scored respiratory events removed (yielding AHI_brain,event-free_ directly). **b**, AHI_GT_ vs. predicted AHI_brain_ in KISS_cpap_, in which respiratory events are suppressed during titration. **c**, AHI_GT_ vs. AHI_brain,event-free_ (Internal Test). In **b**,**c**, dashed lines, identity; red lines and shading, fitted regression with 95% CI.

As a complementary measure of the same discrepancy, we derived AHI_brain,event-free_ by applying the same trained Phase 2 model at inference using only epochs classified as event-free. AHI_brain,event-free_ remained strongly correlated with AHI_GT_ in Internal Test (*r* = 0.77, *p <* 0.001, MAE = 13.26 events/hour; Fig. 2c). The association was more modest but remained significant across the external validation cohorts (SHHS1 *r* = 0.37, SHHS2 *r* = 0.45, and KISS_cpap_*r* = 0.34, all *p <* 0.001; Supplementary Fig. 4b, Supplementary Table 3). These findings suggested that EEG epochs classified as event-free may still contain information related to an individual’s respiratory-event frequency. However, because event-free status was defined at the 30-second epoch level, we next examined whether this association could be explained by the event-labeling criterion itself.

Under our labeling criterion, epochs classified as event-free could still contain less than 10 seconds of respiratory-event overlap. We therefore tested progressively wider exclusion windows around scored events. Such overlap was limited (6.99% of event-free epochs) and fell further to 1.23% with wider exclusion (Supplementary Fig. 5a).

Because respiratory events are often followed by arousals, we also examined the temporal distribution of arousal-related activity. Arousal-positive epochs were concentrated around respiratory events, including the immediately following epoch (Supplementary Fig. 5b). We next examined how progressively removing epochs around scored events affected the association between AHI_brain,event-free_ and AHI_GT_. The correlation weakened as the exclusion window widened but remained evident even under more stringent exclusion (Supplementary Fig. 5c). Within the remaining event-free epochs, both model event probability and false-positive rate rose monotonically with AHI_GT_ severity (Supplementary Fig. 5d,e). Together, these analyses indicate that the severity-related signal in event-free EEG is not explained solely by brief event overlap or nearby post-event activity.

We next examined whether the association between AHI_brain,event-free_ and AHI_GT_ could be explained by differences in available EEG sequence length. Across sequence-construction strategies designed to control for length as a potential confound, a model trained specifically on event-free epochs recovered strong agreement with AHI_GT_ (up to *r* = 0.93; Supplementary Fig. 6), whereas simply shortening the input to a model trained on all epochs degraded performance. These findings indicate that AHI-related information can be learned from event-free EEG content itself and is not attributable to input length alone.

Finally, we examined whether ΔAHI_brain_ and AHI_brain,event-free_ simply captured the same estimation error. The two metrics were moderately related but shared less variance than a length-matched null benchmark (observed *R*^2^ = 0.481 vs. null *R*^2^ = 0.547; Δ*R*^2^ = *−*0.066; Supplementary Fig. 7), indicating that their relationship was not greater than expected from matched input-length reduction alone. Together, these robustness analyses support ΔAHI_brain_ and AHI_brain,event-free_ as overlapping but nonredundant measures of the EEG–respiratory discrepancy.

### Association with sleep macro- and microstructure

Partial correlations (controlling for AHI_GT_, age, BMI, and sex; Benjamini–Hochberg FDR-adjusted *q*-values within each cohort *×* target combination, *q <* 0.05) between ΔAHI_brain_/AHI_brain,event-free_ and a panel of sleep macro-architecture and EEG-microstructure features showed a broadly consistent pattern across Internal, SHHS1, SHHS2, and KISS_cpap_ (Fig. 3). Both metrics were positively associated with lighter, more fragmented sleep macro-architecture (e.g., higher N1%, higher stage-transition and N2-to-N1 transition rates, higher arousal index) and negatively associated with deeper, more consolidated sleep (higher N3%/REM%, higher sleep efficiency, longer TST). Within each cohort, ΔAHI_brain_ and AHI_brain,event-free_ pointed in the same direction for nearly all features, indicating a shared association with sleep fragmentation at the group level despite their nonredundancy at the individual level. Across cohorts, most macro-architecture associations were directionally consistent, although a few features, such as relative theta power in REM, showed opposite directions across cohorts (positive in SHHS1/SHHS2, negative in Internal and KISS_cpap_; Fig. 3). Associations with EEG-microstructure features (relative sigma, delta, and theta power; slow-oscillation and spindle density) were generally weaker and less consistently significant across cohorts than the macro-architecture associations described above.

**Figure 3.**
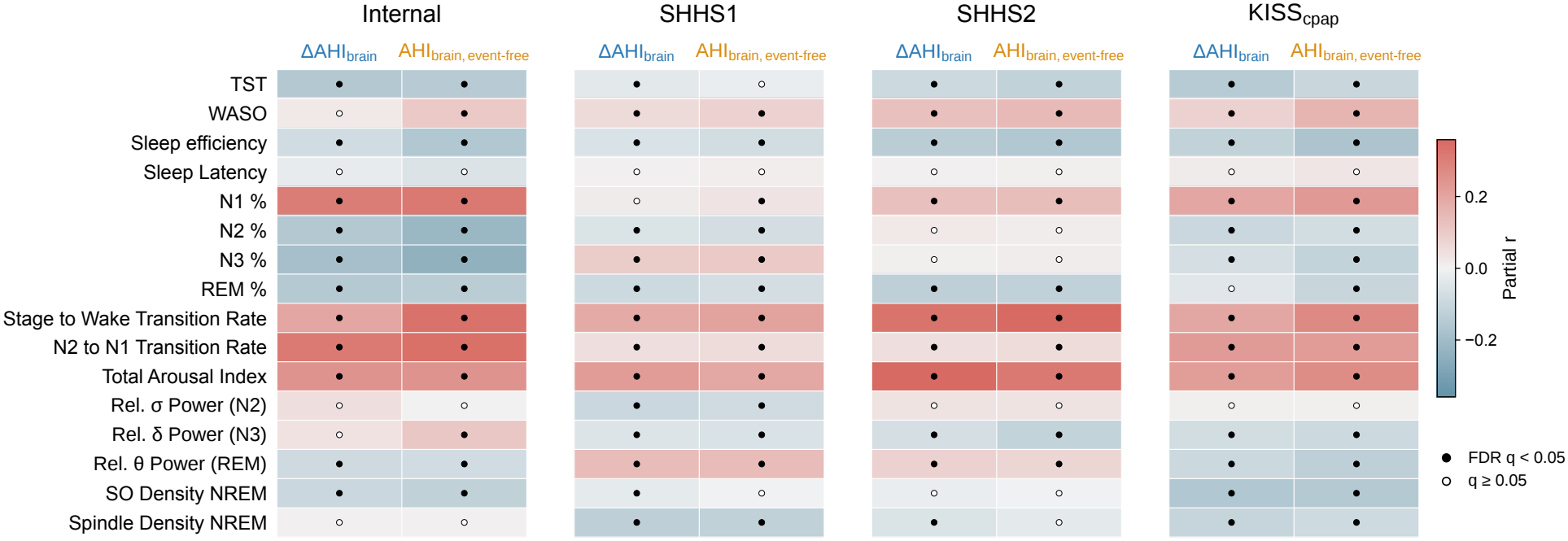
Partial correlations between sleep metrics and ΔAHI_brain_ versus AHI_brain,event-free_. Partial Pearson correlations between ΔAHI_brain_ or AHI_brain,event-free_ and sleep macro- and microstructure metrics, adjusted for AHI_GT_, age, BMI, and sex, shown as a heatmap (color scale, partial *r*) across Internal, SHHS1, SHHS2, and KISS_cpap_. Filled circles, *q <* 0.05; open circles, not significant.

### EEG correlates of predicted respiratory-event probability

As an initial visualization of the Phase 1 representations, we projected the patient-level representations into a low-dimensional embedding and examined their organization with respect to conventional AHI severity and the two EEG-derived metrics. Embeddings derived from both all epochs and event-free epochs showed continuous variation with AHI_GT_ severity across all four cohorts (Supplementary Fig. 8). Within individual AHI_GT_ severity strata, the all-epoch embedding also showed variation associated with ΔAHI_brain_, whereas the event-free embedding showed variation associated with AHI_brain,event-free_ (Supplementary Fig. 9). These analyses were descriptive and were used to visualize the organization of the Phase 1 representations.

To relate the Phase 1 representations to interpretable EEG physiology, we next examined whether the model’s predicted event probability was systematically related to underlying EEG characteristics rather than merely reproducing the respiratory-event label. From these features, two representative features per sleep stage were selected for subsequent analyses based on their consistency across cohorts (see Supplementary Note 2 for the selection procedure): relative delta power and relative fast-delta power in N1, theta/alpha ratio and slow-oscillation amplitude in N2, relative slow-delta power and relative theta power in N3, and relative alpha power and relative delta power in REM. For each feature, we plotted its standardized effect size relative to TN epochs against the model’s predicted event probability. We examined this relationship separately for event and non-event epochs across four cohorts (Internal, SHHS1, SHHS2, and KISS_cpap_; Fig. 4a). The full feature results are shown by sleep stage in Supplementary Figs. 10–13.

**Figure 4.**
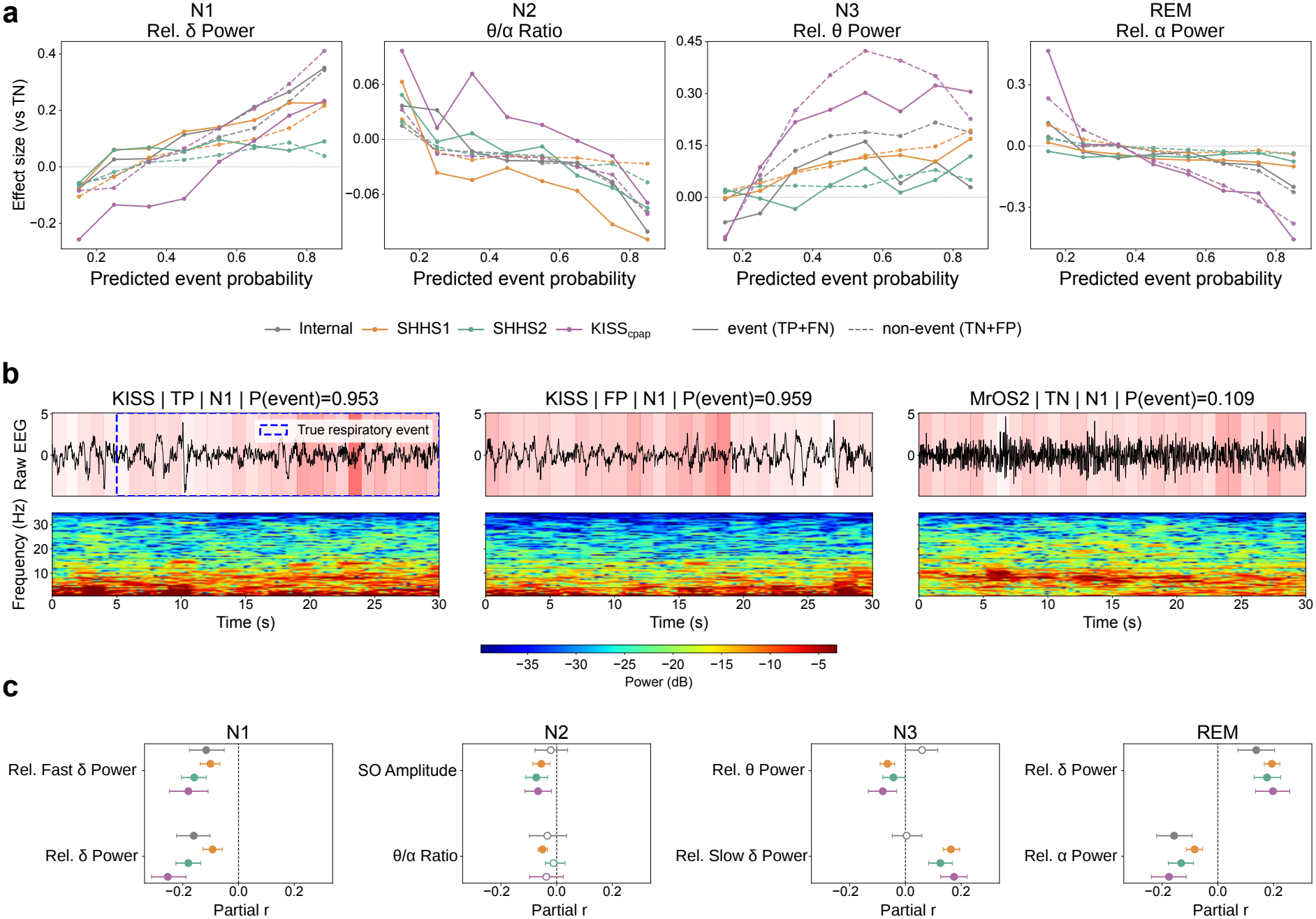
Epoch-level construct validity of ΔAHI_brain_ and AHI_brain,event-free_. **a**, Dose-response of one representative EEG feature per sleep stage (N1, relative delta power; N2, theta/alpha ratio; N3, relative theta power; REM, relative alpha power); solid lines, event (TP+FN) epochs; dashed lines, non-event (TN+FP) epochs. The full eight-feature panel is shown per stage in Supplementary Figs. 10–13. **b**, Attention-overlay examples (N1 sleep). Raw EEG (top) and spectrogram (bottom) with Phase 1 attention weight overlaid as shading; blue dashed box, scored respiratory event. Left, true-positive epoch; middle, false-positive epoch; right, true-negative epoch. **c**, Bridging epoch-level features to patient-level metrics (Connection 1): partial *r* (AHI_GT_-adjusted) between a patient’s TN-epoch mean feature value and AHI_brain,event-free_. Filled circles, 95% CI excludes 0. Connection 2 (feature-deviation slope vs. ΔAHI_brain_) is reported separately in Supplementary Fig. 14.

For most features, EEG characteristics changed systematically with predicted event probability in both respiratory-event and non-event epochs. At higher predicted probabilities, the two groups tended to converge toward similar feature values. Thus, EEG feature values varied with the model’s predicted event probability even when the respiratory-event labels differed. This pattern was also evident in discordant epochs, suggesting that high-confidence predictions in non-event epochs were not simply unrelated classification errors but were associated with EEG characteristics similar to those accompanying high event probability in event epochs.

Figure 4b illustrates this relationship in three representative N1 epochs spanning different levels of agreement between the respiratory-event label and model prediction. An event epoch with high predicted probability (*P* = 0.953; TP) and a non-event epoch with similarly high predicted probability (*P* = 0.959; FP) are contrasted with a non-event epoch with low predicted probability (*P* = 0.109; TN). Both high-probability examples showed increased delta-band power in regions receiving greater attention, consistent with the positive relationship between predicted probability and relative delta power in N1 observed in Fig. 4a. Notably, this pattern was also present in the FP epoch despite its negative respiratory-event label. In contrast, the low-probability TN epoch showed relatively stronger alpha-band activity together with a more broadly distributed attention pattern. These examples show, at the level of individual epochs, the same probability-dependent EEG patterns observed at the cohort level.

### Bridging epoch-level features to patient-level metrics

The preceding analysis (Fig. 4a) showed that EEG feature values varied systematically with the model’s predicted event probability at the epoch level. However, a pattern observed across pooled epochs does not necessarily explain inter-individual differences in patient-level ΔAHI_brain_ and AHI_brain,event-free_. We therefore asked whether the same EEG features could bridge these two levels of analysis. We defined each patient’s mean feature value across true-negative (TN) epochs as their baseline and performed two complementary analyses using the selected eight-feature panel (precise definitions in Methods).

Connection 1 tested whether inter-individual differences in these TN baseline values were associated with AHI_brain,event-free_ after adjustment for AHI_GT_ (Fig. 4c). Several features, including REM relative alpha and delta power and N1 relative delta and fast-delta power, showed directionally consistent and statistically significant associations across all four cohorts. Other features, including N2 theta/alpha ratio, N2 slow-oscillation amplitude, and N3 relative slow-delta power, showed weaker or cohort-inconsistent associations.

Overall, Connection 1 showed that patient-specific baseline EEG characteristics during TN epochs were systematically related to AHI_brain,event-free_ independently of AHI_GT_.

Connection 2 tested whether patient-specific EEG reactivity was associated with ΔAHI_brain_. Using each patient’s TN mean as the baseline, we quantified how strongly feature values deviated from this baseline as predicted event probability increased. These associations were weaker and less consistent, with no feature showing consistent replication across the three diagnostic cohorts (Supplementary Fig. 14).

### Mortality associations beyond AHI

We next asked whether ΔAHI_brain_ and AHI_brain,event-free_ were also associated with long-term mortality. We restricted this analysis to participants without CVD at baseline to evaluate prognostic associations in the absence of pre-existing cardio-vascular disease. We examined associations with all-cause, CVD, and non-CVD mortality, and whether these associations persisted after accounting for AHI_GT_ and standard clinical risk factors—age, sex, BMI, race, smoking, diabetes, hypertension, and dyslipidemia. The baseline CVD-free SHHS dataset included 4,694 participants with complete covariate data, with 827 all-cause, 181 CVD, and 646 non-CVD deaths. Associations were evaluated using progressively adjusted Cox proportional-hazards models (Supplementary Tables 6–7).

Kaplan–Meier curves showed survival differences across quartiles of both EEG-derived metrics (Fig. 5). For ΔAHI_brain_, quartile differences were significant for all-cause and non-CVD mortality (both log-rank *p <* 0.001), whereas the difference for CVD mortality did not reach statistical significance (*p* = 0.052; Fig. 5a–c). For AHI_brain,event-free_, quartile differences were significant for all three outcomes (all log-rank *p <* 0.001; Fig. 5d–f).

**Figure 5.**
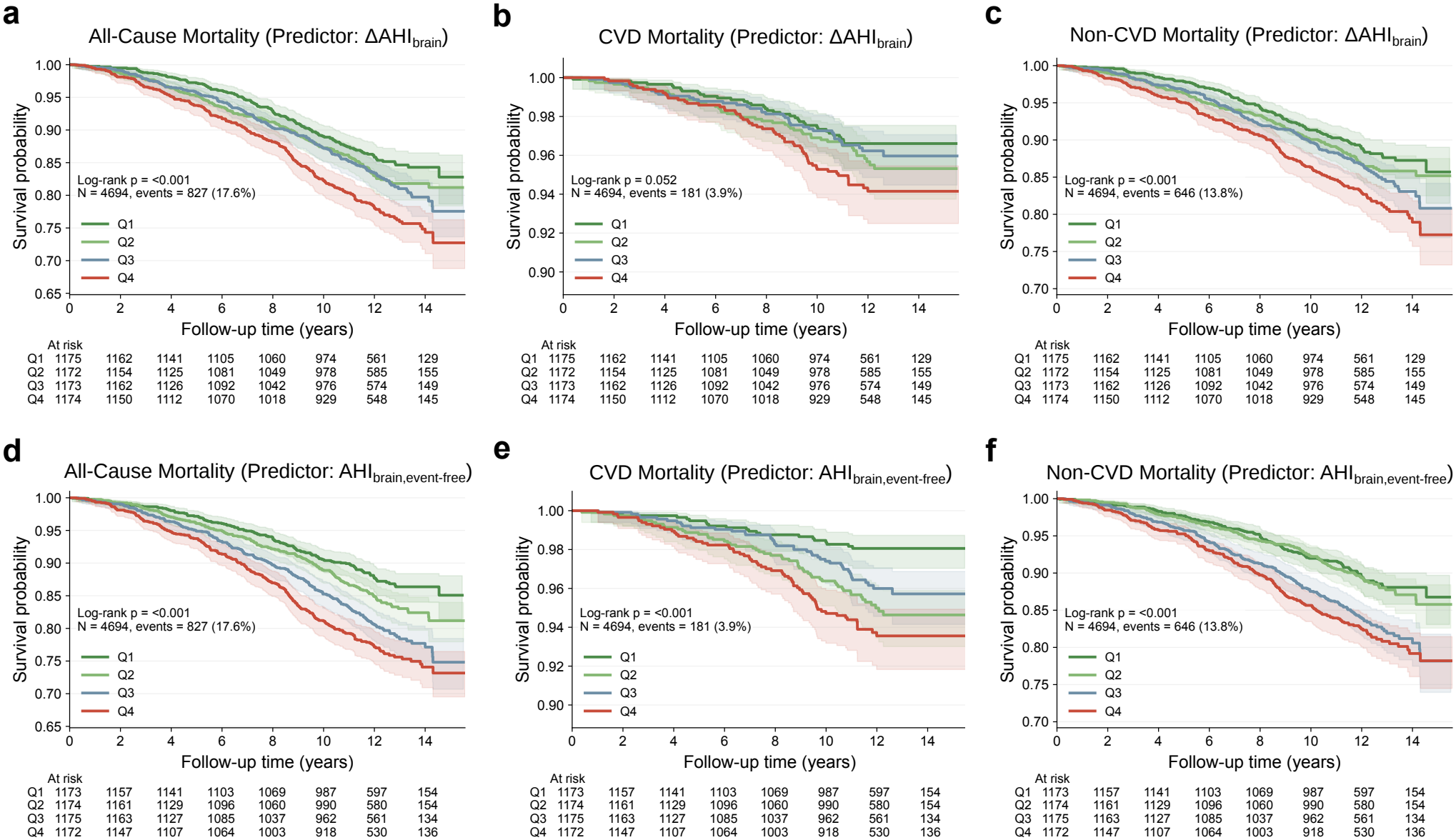
Kaplan–Meier survival curves by predictor quartile (Q1–Q4) in the baseline CVD-free SHHS cohort. **a–c**, All-cause (**a**), CVD (**b**), and non-CVD (**c**) mortality by ΔAHI_brain_ quartile. **d–f**, All-cause (**d**), CVD (**e**), and non-CVD (**f**) mortality by AHI_brain,event-free_ quartile. Log-rank *p* values were *<* 0.001, 0.052, and *<* 0.001 for **a–c**, respectively, and *<* 0.001 for **d–f**. Shaded bands indicate 95% CIs; numbers at risk are shown below each plot.

We first evaluated AHI_GT_, ΔAHI_brain_, and AHI_brain,event-free_ in separate Cox models, each adjusted for the same clinical covariates (Fig. 6a; Supplementary Tables 4–5). After full adjustment, ΔAHI_brain_ and AHI_brain,event-free_ remained significantly associated with all-cause mortality, whereas AHI_GT_ did not. For CVD mortality, none of the three predictors showed a significant per-SD association. For non-CVD mortality, all three predictors were significantly associated with risk, with similar effect sizes across AHI_GT_, ΔAHI_brain_, and AHI_brain,event-free_.

**Figure 6.**
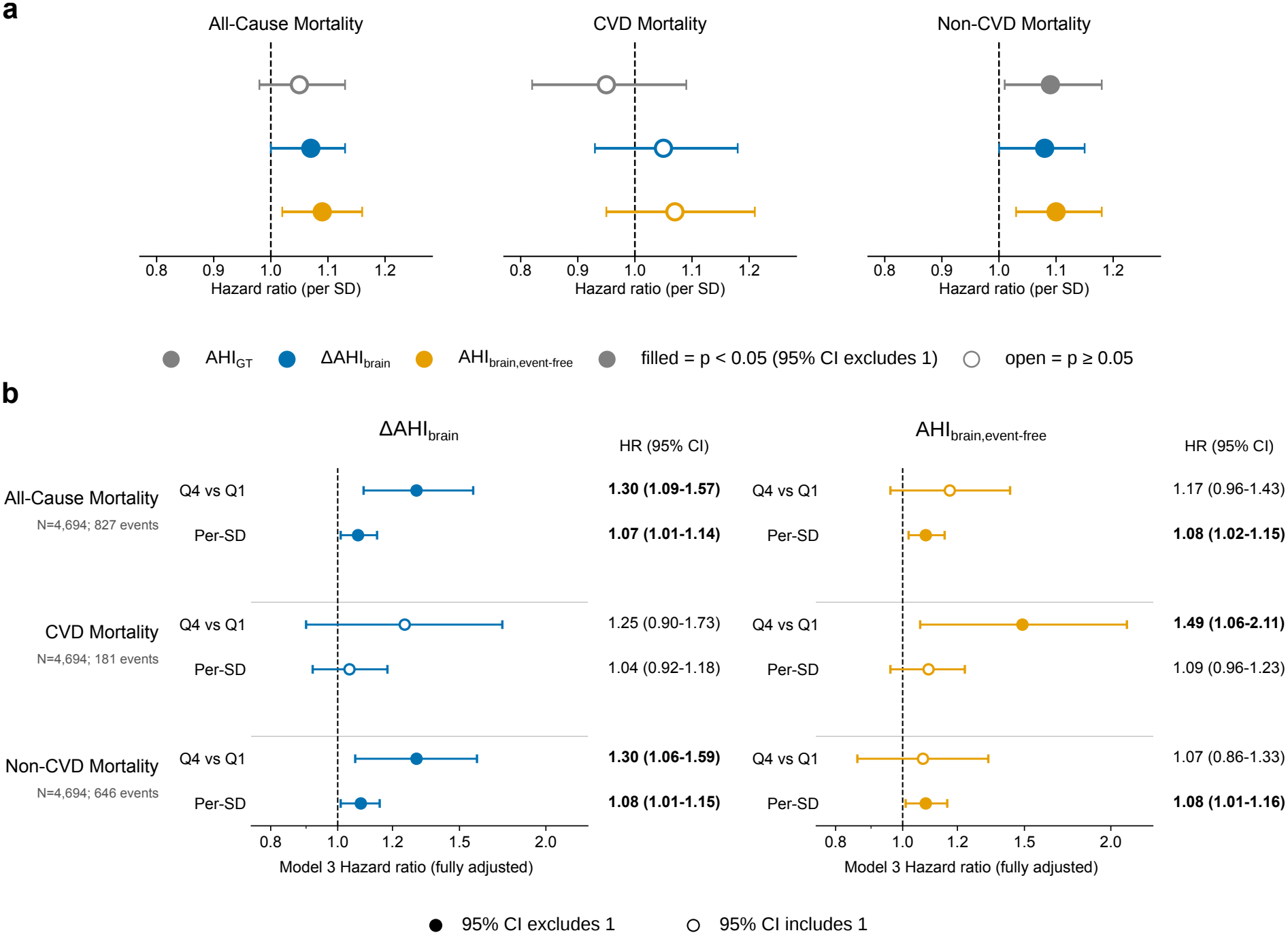
**a**, Head-to-head comparison of per-SD hazard ratios for AHI_GT_, ΔAHI_brain_, and AHI_brain,event-free_ against all-cause, CVD, and non-CVD mortality, each fit independently in fully adjusted Cox models (Model 3: age, sex, BMI, race, smoking status, diabetes, hypertension, dyslipidemia); the three predictors were not included together in the same model. **b**, Forest plot of Model 3 (fully adjusted) hazard ratios for ΔAHI_brain_ and AHI_brain,event-free_ against all-cause, CVD, and non-CVD mortality, shown for both extreme-quartile (Q4 vs. Q1) and continuous per-SD parameterizations. In **a**,**b**, filled markers indicate hazard ratios whose 95% CI excludes 1; open markers indicate CIs that include 1.

We next examined whether the associations of the two EEG-derived metrics persisted after adjustment for AHI_GT_ and standard clinical covariates (Fig. 6b). Higher ΔAHI_brain_ was associated with all-cause mortality in both the quartile (Q4 vs. Q1: HR 1.30, 95% CI 1.09–1.57) and continuous analyses (per SD: HR 1.07, 95% CI 1.01–1.14). Similar associations were observed for non-CVD mortality (Q4 vs. Q1: HR 1.30, 95% CI 1.06–1.59; per SD: HR 1.08, 95% CI 1.01–1.15). No significant association was observed with CVD mortality.

AHI_brain,event-free_ was also associated with all-cause and non-CVD mortality in the fully adjusted models. The per-SD HRs were 1.08 (95% CI 1.02–1.15) for all-cause mortality and 1.08 (95% CI 1.01–1.16) for non-CVD mortality. For CVD mortality, the Q4-versus-Q1 association was statistically significant (HR 1.49, 95% CI 1.06–2.11). ΔAHI_brain_ and AHI_brain,event-free_ were both associated with all-cause and non-CVD mortality, with comparable per-SD HRs for both outcomes. For CVD mortality, a significant association was observed only for the quartile parameterization of AHI_brain,event-free_.

The addition of either EEG-derived metric (ΔAHI_brain_ or AHI_brain,event-free_) to a model adjusted for AHI_GT_ and clinical covariates resulted in only minimal changes in model discrimination (ΔC-index *<* 0.001; Supplementary Tables 6–7). The proportional-hazards assumption was satisfied for all reported models.

## Discussion

This study asked what information the AHI may miss. AHI quantifies respiratory events but treats events as equivalent regardless of their physiological consequence for an individual. We trained a deep-learning model to estimate AHI from a single EEG channel and validated it in an internal test set and three external test datasets. We then examined whether the discrepancy between EEG-derived and conventionally scored AHI carries meaningful information beyond residual model error. We characterized this EEG–Respiratory Gap using two complementary metrics. ΔAHI_brain_ is the residual of the full-recording model output after accounting for AHI_GT_ and is therefore linearly independent of AHI_GT_ by construction. In contrast, AHI_brain,event-free_ is derived from epochs without scored respiratory events and remains correlated with AHI_GT_. Both metrics were associated with epoch-level EEG features and sleep architecture in physiologically consistent directions. Among participants free of cardiovascular disease at base-line, the two metrics showed broadly similar associations with all-cause and non-CVD mortality; AHI_brain,event-free_ was additionally associated with CVD mortality in the highest-versus-lowest quartile comparison when adjusted for conventionally scored AHI and standard clinical covariates.

A natural concern is that the EEG–Respiratory Gap may reflect imperfect AHI estimation rather than meaningful inter-individual variation. ΔAHI_brain_ was designed to quantify this discrepancy while removing its linear dependence on conventionally scored AHI. Specifically, it represents how much a patient’s AHI_brain_ is higher or lower than expected for their AHI_GT_, rather than the simple prediction error between the two values. Whether this residual variation is meaningful must therefore be established through its associations with independent physiological and clinical outcomes.

AHI_brain,event-free_ provides a complementary perspective by estimating AHI_brain_ using only epochs classified as non-event. This metric should not be interpreted as completely independent of respiratory events. Brief event overlap can remain because of the epoch-level labeling criterion, and physiologically meaningful responses may extend beyond the event-positive epoch itself. However, such overlap was limited, and AHI-related patterns persisted when progressively wider regions surrounding respiratory events were excluded (Supplementary Fig. 5a,c–e). Arousal activity also remained elevated immediately after respiratory events (Supplementary Fig. 5b), suggesting that event-related EEG responses can extend beyond the labeled event epoch. We therefore interpret AHI_brain,event-free_ as capturing EEG-derived respiratory information present outside epochs classified as respiratory-event, rather than as a purely event-independent signal. Additional controls, including models trained using event-free epochs alone, further showed that AHI-related information could be learned from the retained EEG itself and was not explained by input-length differences alone (Supplementary Fig. 6).

The physiological and prognostic associations of these metrics provide further evidence that the EEG–Respiratory Gap is not simply uninformative estimation noise. Both metrics were associated with sleep macrostructure and, less consistently, EEG microstructure after adjustment for AHI_GT_. Epoch-level analyses further showed systematic EEG feature changes with predicted respiratory-event probability, including in non-event epochs. At the patient level, these EEG characteristics were more consistently linked to AHI_brain,event-free_, whereas the corresponding feature-level associations with ΔAHI_brain_ were weaker. Finally, both ΔAHI_brain_ and AHI_brain,event-free_ remained associated with all-cause and non-CVD mortality after adjustment for AHI_GT_ and standard clinical covariates. Together, these findings support the interpretation of the two metrics as complementary measures of EEG–Respiratory discrepancy rather than simple measures of AHI prediction error.

The clearest epoch-level substrate for AHI_brain,event-free_ came from a patient’s own baseline EEG state across sleep stages. The direction of these associations formed a coherent rather than a scattered pattern. Higher AHI_brain,event-free_ was consistently linked to lower delta-band power at a patient’s own baseline during epochs without scored respiratory events: lower relative delta and fast-delta power in N1, lower relative slow-delta power in N3, and lower relative delta power in REM. At the same time, higher AHI_brain,event-free_ was associated with greater faster-rhythm activity, specifically higher relative theta power in N3 and higher relative alpha power in REM. Together, these findings describe a baseline EEG shifted away from slow, high-amplitude activity and toward faster rhythms across multiple stages. This pattern was most marked in REM, where elevated alpha content is itself atypical. REM sleep is normally characterized by low-voltage, desynchronized cortical activity. Alpha intrusion into REM, together with reduced REM delta, is therefore more consistent with a persistently lighter, more wake-adjacent REM state than with ordinary REM physiology. The prominence of this pattern in REM may also be relevant to cardiovascular physiology. Independent of overall AHI, REM sleep carries the greatest autonomic and hemodynamic instability of the night, and respiratory events concentrated in REM are an established predictor of hypertension and impaired nocturnal blood-pressure dipping^35,36^. A baseline tendency toward a lighter, more arousable REM state, of the kind captured here, may therefore reflect autonomic vulnerability relevant to cardiovascular risk. Consistent with this possibility, AHI_brain,event-free_ was the only one of the two gap metrics to show an association with CVD mortality, although this was limited to the highest-versus-lowest quartile comparison and was not significant in the continuous analysis. Severity metrics beyond simple event counting, such as hypoxic-burden-based measures, have like-wise been associated with cardiovascular mortality^7,8^. This interpretation is also compatible with the observation that AHI_brain,event-free_ remained elevated in KISS_cpap_ even though scored respiratory events were actively suppressed by titration. Patients undergo CPAP titration because they already carry a clinical diagnosis of significant OSA. A single night of pressure normalization can suppress discrete, fully scored events without necessarily eliminating more subtle flow limitation or reversing EEG changes that may have accumulated over a longer period of untreated disease. EEG alterations of this kind could therefore persist during a titration night even as the scored event burden is substantially reduced.

In contrast, ΔAHI_brain_ was associated with all-cause and non-CVD mortality but not CVD mortality. Non-CVD mortality encompasses multiple causes, including cancer, respiratory disease, and neurodegenerative conditions, making this association difficult to attribute to a specific physiological pathway. To explore a potential physiological correlate of ΔAHI_brain_, we examined whether it might relate to inter-individual differences in EEG reactivity to model-predicted respiratory events, including arousal-related responses. However, the corresponding feature-reactivity analysis showed limited consistency across cohorts (Supplementary Fig. 14), and respiratory arousal threshold was not measured directly in this study. A reactivity-based interpretation of ΔAHI_brain_ therefore remains a hypothesis requiring further investigation.

Our metrics are conceptually related to, but distinct from, other EEG/PSG-derived measures relevant to OSA. Arousal burden, defined as the cumulative duration of scored arousals normalized to total sleep time, has been associated with long-term all-cause and cardiovascular mortality in MrOS and SHHS^15^, both of which were included in the present study. However, a separate analysis found no significant associations of arousal burden with cardiovascular outcomes or all-cause mortality in MESA, while in MrOS an association was observed for cardiovascular mortality but not for all-cause mortality or incident CVD^37^. Overall, these findings suggest that the prognostic associations of arousal burden may vary across cohorts and outcomes. In addition, arousal burden is defined directly from scored arousal events and does not distinguish respiratory from non-respiratory arousals. In contrast, ΔAHI_brain_ quantifies the discrepancy between EEG-estimated and measured AHI after removing its systematic dependence on AHI_GT_, whereas AHI_brain,event-free_ is estimated specifically from epochs without scored respiratory events. Continuous EEG-derived measures such as the odds ratio product^38^ similarly characterize aspects of cortical state and sleep depth, but were not designed to estimate AHI or its discrepancy from conventionally scored values. The extent to which ΔAHI_brain_ and AHI_brain,event-free_ overlap with arousal burden, the odds ratio product, or other established EEG-derived measures remains to be determined.

More broadly, the sleep-microstructure analyses showed that spindle and slow-oscillation density were associated with both discrepancy metrics, alongside the macro-architectural findings described above. These features have been implicated in restorative functions of NREM sleep, including memory consolidation^39^, synaptic homeostasis^40^, and processes related to glymphatic clearance^41^. Their associations with the discrepancy metrics suggest that the physiological information reflected by the EEG–Respiratory Gap may extend beyond cardiovascular processes and may also be relevant to broader health outcomes. Recent work in the same MrOS and SHHS cohorts similarly showed that deep-learning-derived EEG power-band features were independently associated with all-cause mortality^42^, providing complementary evidence for the broader prognostic relevance of EEG-derived measures.

Taken together, the associations with sleep physiology and long-term outcomes suggest that the EEG–Respiratory Gap may provide information complementary to conventionally scored AHI. Both metrics remained associated with all-cause and non-CVD mortality after adjustment for AHI_GT_ and standard clinical risk factors. From a practical standpoint, they are derived from EEG already recorded during standard overnight polysomnography, and therefore require no additional sensors or patient burden. Their clinical utility, however, will require prospective validation and evaluation of whether they provide actionable information beyond established risk factors.

Several limitations should be considered. First, this observational study establishes association rather than causation, and the physiological pathways linking the EEG–Respiratory Gap to mortality were not directly tested. Key measures relevant to our physiological interpretations, including arousal threshold, blood pressure, and heart-rate variability, were not directly assessed. Spindle and slow-oscillation features were also not evaluated in relation to mortality, and outcome specificity was not formally tested using interaction analyses. Second, some EEG associations varied across cohorts. For example, relative theta power in REM showed opposite associations with the discrepancy metrics in SHHS compared with our internal and KISS_cpap_ cohorts. Such heterogeneity may reflect differences in study populations, recording devices, or scoring criteria and warrants caution when interpreting individual EEG features across cohorts. Third, the survival analyses were limited to SHHS, an older community-dwelling cohort without CVD at baseline, which may limit generalizability to other populations. Non-CVD mortality was a heterogeneous residual category comprising deaths not classified as CVD mortality and was not further decomposed by underlying cause; the specific conditions driving this association therefore remain to be identified. KISS_cpap_ could not be included in the survival analyses because of its titration-night design. Finally, computing the metrics currently requires a full overnight EEG recording and inference through a trained model. Clinical implementation will therefore require prospective validation and workflow integration. Cognitive and neurodegenerative outcomes were beyond the scope of this study and are being investigated separately.

In summary, ΔAHI_brain_ and AHI_brain,event-free_ provide complementary measures of the discrepancy between EEG-derived and conventionally measured AHI. Their associations with sleep architecture, EEG characteristics, and long-term mortality suggest that this discrepancy contains structured information beyond simple AHI prediction error. These findings support the EEG–Respiratory Gap as an interpretable complement to conventional AHI and suggest that EEG already collected during polysomnography may contain clinically relevant information not captured by standard respiratory-event frequency alone.

## Methods

### Data sources and ethics

Five polysomnography (PSG) datasets were used. KISS is an internal, multi-cohort dataset collected across four institutions, comprising two subsets: a diagnostic-night PSG cohort (KISS_diag_) and a CPAP titration-night PSG cohort (KISS_cpap_). The remaining three datasets – MESA^43,44^, MrOS^43,45^ (visits 1 and 2), and SHHS^43,46^ (visits 1 and 2) – are open datasets available through the National Sleep Research Resource (NSRR). Detailed dataset descriptions are provided in Supplementary Note 1.

KISS_cpap_ consists of single-night titration studies in which airway pressure is adjusted in real time to suppress respiratory events. Its within-night AHI_GT_ distribution is therefore markedly compressed relative to the diagnostic-night cohorts (Supplementary Table 1), and KISS_cpap_ was treated separately when interpreting model performance and downstream analyses.

This study – including analysis of KISS together with the publicly available MESA, MrOS, and SHHS datasets – was approved by the responsible Institutional Review Board (IRB approval number: E-2510-019-1681), which covers large-scale secondary analysis across these datasets rather than the original collection of KISS itself. The original collection of KISS was separately approved by the institutional review boards of Seoul National University Hospital (C-2007-179-1143) and Chuncheon Sacred Heart Hospital, Hallym University College of Medicine (2021-03-005), with informed consent waived because the data were de-identified^47^. Use of MESA, MrOS, and SHHS data was additionally conducted under the data use agreements of the respective parent studies, each of which obtained institutional review board approval and participant informed consent at the time of original data collection^43^. All data were de-identified prior to analysis.

Per-cohort baseline characteristics are reported in Supplementary Table 1.

### Data preprocessing

A single central EEG derivation (C4-A1), among the channels commonly used in sleep research^48^, was used as model input for every cohort. Raw single-channel EEG was preprocessed identically across datasets. A 60 Hz notch filter was applied to remove line noise, followed by a 0.5–35 Hz band-pass filter (8th-order Butterworth, zero-phase), and the signal was resampled to 128 Hz. The filtered signal was decomposed into five sub-bands (0.5–4, 4–8, 8–12, 12–15, and 15–35 Hz), stacked, and z-score normalized per recording before being used as model input. Recordings were trimmed at the sleep-period boundary by removing wake periods exceeding 30 minutes immediately before sleep onset or after sleep offset, retaining the sleep period plus up to 30 minutes of adjacent wake. Signals were then segmented into consecutive, non-overlapping 30-second epochs for sleep-stage and respiratory-event labeling and prediction.

Sleep-stage scoring conventions differed across datasets. KISS was scored using AASM criteria, whereas MESA, MrOS, and SHHS were originally scored using Rechtschaffen & Kales (R&K) criteria. To harmonize sleep-stage labels, R&K stages 3 and 4 were merged into a single N3 stage, matching the five-class scheme (Wake, N1, N2, N3, REM) used throughout this study. Sleep-stage ground truth was otherwise taken directly from each cohort’s native per-epoch scoring.

Ground-truth AHI (AHI_GT_) reflects the AASM 2012 alternative hypopnea rule, requiring either *≥*3% oxygen desaturation or an arousal, in each dataset’s official derived-variables file. However, individual hypopnea annotations in SHHS, MrOS, and MESA were originally scored by flow reduction alone without this requirement. Event-level hypopnea annotations in these cohorts were therefore re-scored according to AASM 2012 criteria using a standardized annotation-refinement pipeline^49^; KISS already followed AASM criteria at the event level. An epoch was labeled as an event epoch if apnea or hypopnea occupied at least 10 seconds of the 30-second epoch; otherwise, it was labeled as a non-event epoch. This criterion was applied identically across all five datasets.

### Model architecture

brAHInet (Fig. 1b) comprises two stages. Phase 1 is a multi-task deep neural network operating on each 30-second central (C4-A1) EEG epoch. The five-band-filtered signal forms an input tensor *X* ∈ ℝ ^5×3840^ (5 frequency bands × 128 Hz × 30 s). *X* is encoded by a ModernTCN backbone^34^, a pure-convolutional time-series architecture that alternates depth-wise temporal convolutions with pointwise cross-channel convolutions across successive downsampling stages, yielding a feature map *F* ∈ ℝ^*M×D×N*^ (*M*=5 frequency bands, *D* feature dimension, *N* temporal patches). A self-attention block applies multi-head self-attention^50,51^ over *F* (reshaped to *N* × *MD*) with a residual connection, giving updated features *Z*. A sleep-staging head maps *Z* to per-epoch stage logits *s* ∈ ℝ5 (Wake/N1/N2/N3/REM). An additive-attention module then combines *Z* with softmax(*s*) as context, allowing the respiratory-event head to incorporate the model’s sleep-stage prediction. The event head outputs binary event logits *e* ∈ ℝ ^2^, corresponding to event and non-event classes, together with an intermediate event representation *ϕ* ∈ ℝ ^30×80^ for each epoch. Here, the 30 positions correspond to one-second temporal representations within the 30-second epoch, and 80 is the hidden feature dimension of the event head. Phase 1 is trained with a combined loss *L* = *L* _stage_ + *L*_event_, where *L*_stage_ is cross-entropy on the five-class stage label and _event_ is class-weighted cross-entropy on the binary event label. A detailed layer-by-layer diagram, including tensor shapes, is provided in Supplementary Fig. 2.

Phase 2 predicts patient-level AHI_brain_ from the temporally ordered sequence of Phase 1 event representations. For a patient with *T* epochs, the representations are assembled as *X*_*p*_ = [*φ*_1_, …, *φ*_*T*_ ], where each *φ*_*t*_ *∈* ℝ^30*×*80^. Each epoch representation is flattened to a 2,400-dimensional vector and linearly projected to a 256-dimensional embedding. Sinusoidal positional encoding is added, and a learnable [CLS] token is prepended before the sequence is passed through a three-layer Transformer encoder^50^ with four attention heads and dropout of 0.1. The encoded [CLS] representation is layer-normalized and passed through a linear regression head to obtain the patient-level prediction. Phase 2 is trained to predict log(1 + AHI_GT_) using a weighted mean absolute error loss that assigns greater weight to the lower and higher ends of the AHI distribution, with the model output transformed back to the AHI scale for evaluation.

Phase 1 was optimized using Adam with a learning rate of 1 *×*10^*−*3^ and a batch size of 512. Training was performed for up to 30 epochs with early stopping (patience 7) based on validation performance across the respiratory-event and sleep-stage tasks. Phase 2 was optimized using AdamW with a learning rate of approximately 2 *×*10^*−*5^, weight decay of 0.01, and a batch size of 1. Training was performed for up to 50 epochs with early stopping (patience 10), with gradient clipping at a maximum norm of 4.0. The checkpoint with the best validation performance was retained for subsequent analyses.

Phase 1 and Phase 2 models were implemented in Python 3.11.13 using PyTorch 2.5.1 (CUDA 12.1 build; cuDNN 9.1.0) and scikit-learn 1.7.1, with h5py 3.14.0 used for EEG data I/O and Weights & Biases 0.19.6 for experiment tracking. Each training run used a single NVIDIA RTX A6000 GPU (49 GB VRAM) on a Linux (Ubuntu 22.04.3 LTS) server with 128 CPU cores and 540 GB RAM.

For comparison, EEG-CLNet^21^, a baseline convolutional model that performs sleep staging and respiratory-event detection via a shared convolutional feature extractor with cross-task knowledge distillation between the two tasks, was trained and evaluated on the identical data splits as a comparator for both epoch-level tasks (Table 2). EEG-CLNet does not include a patient-level AHI-regression component.

### Bias correction and null-benchmark control for ΔAHI_brain_ and AHI_brain,event-free_

ΔAHI_brain_ is the residual of an OLS regression of AHI_brain_ on AHI_GT_. For Internal Test, the regression’s slope and intercept were fit on the held-out Validation partition and applied to Internal Test, so that ΔAHI_brain_’s orthogonality to AHI_GT_ there reflects an out-of-sample correction rather than a trivial consequence of fitting and evaluating on the same data, consistent with recommended procedures for correcting regression-based prediction bias in similar predicted-observed discrepancy frameworks^52,53^. For each External Test cohort (SHHS1, SHHS2, KISS_cpap_), no comparable held-out validation subset exists, so the regression was instead fit and applied within that same cohort.

To distinguish the effect of selectively removing respiratory-event epochs from that of simply reducing the amount of EEG input, we constructed a length-matched null control. For each patient, we first calculated the proportion of epochs retained in the event-free analysis. We then selected the closest matching condition from the input-length reduction analysis (Supplementary Fig. 6a), in which a similar proportion of the recording was retained without selectively removing respiratory-event epochs. The resulting control prediction was residualized against AHI_GT_ in the same manner as AHI_brain,event-free_. We compared the AHI_GT_-adjusted associations of ΔAHI_brain_ with AHI_brain,event-free_ and with this length-matched control using five-fold cross-validated ridge-regression *R*^2^. Differences in *R*^2^ and 95% confidence intervals were estimated using 1,000 bootstrap samples (seed= 0).

### EEG feature extraction

Candidate EEG microstructure features were extracted using the YASA toolbox (v0.6.5)^54^, applied to the same C4-A1 derivation used by the Phase 1 model. Relative spectral power (delta, slow-delta, fast-delta, theta, alpha, sigma, beta bands) and the theta/alpha ratio were computed directly per 30-second epoch (N1, N2, N3, REM). Sleep spindles and slow oscillations were instead detected on the continuous whole-night signal (YASA’s default detectors), and each detected event was assigned to the 30-second epoch containing its onset, from which epoch- and stage-level (N2, N3) density and amplitude were derived.

### Epoch-level feature analysis

Epoch classification, feature panel construction, person-mean-centering, dose-response binning, monotonicity-based feature selection, and the Connection 1/2 and FDR procedures were performed as described below. Extended cohort-specific analyses and complete candidate-feature results are provided in Supplementary Note 2.

#### Dose-response binning

For each of the four sleep stages, model-predicted event probability was divided into ten bins; the two most extreme bins (nominal centers 0.05 and 0.95) were excluded to avoid instability from sparsely populated tails, leaving eight analysis bins. For each patient and sleep stage, the mean feature value across true-negative (TN) epochs was defined as the patient-specific baseline (TN anchor). Each epoch’s feature value was expressed as a deviation from this baseline^55^, thereby emphasizing within-patient changes rather than differences in overall EEG level between patients. Within each probability bin, these deviations were standardized to obtain the effect sizes shown in Fig. 4a. Within each stage, the two features with the highest monotonicity (rank correlation between probability bin and centered effect size), among those showing a consistent direction across cohorts, were retained for downstream analyses: N1 – relative delta power and relative fast-delta power; N2 – theta/alpha ratio and slow-oscillation amplitude; N3 – relative slow-delta power and relative theta power; REM – relative alpha power and relative delta power.

#### Connection 1 (TN-anchored patient mean vs. AHI_brain,event-free_)

Connection 1 tested whether the patient-specific TN baseline for each retained feature was associated with AHI_brain,event-free_ after controlling for AHI_GT_. Partial Pearson correlations were computed separately for each feature and cohort among patients with at least 10 TN epochs in the relevant sleep stage.

#### Connection 2 (TP+FP deviation slope vs. ΔAHI_brain_)

Connection 2 tested whether patient-specific EEG feature reactivity was associated with ΔAHI_brain_. For each patient, epochs with model-predicted event probability *>* 0.5 (TP+FP) were used to quantify feature deviations from the same TN baseline. Patient-specific reactivity was estimated as the Theil–Sen slope^56^ of feature deviation against predicted event probability, restricted to patients with at least 10 such epochs and at least two distinct probability values. The resulting slopes were correlated with ΔAHI_brain_ separately for each feature and cohort.

#### Epoch embedding analysis

For each patient with *T* epochs, low-dimensional UMAP embeddings were computed from the Phase 1 event-detection representations of shape (*T*, 30, 80), corresponding to 30 one-second sub-epoch positions *×*80-dimensional feature vectors. Separate embeddings were constructed using all epochs and event-free epochs to examine whether the learned representations contained structure related to AHI_GT_, ΔAHI_brain_, and AHI_brain,event-free_.

### Statistical analysis

All statistical analyses were performed in Python 3.11.13 (pandas 2.3.1, numpy 1.26.4, scipy 1.16.0, statsmodels 0.14.5, pingouin 0.6.1, lifelines 0.30.3, matplotlib 3.10.0). Unless otherwise noted, correlation and regression statistics used two-tailed tests at *α* = 0.05 (or BH-FDR-corrected *q <* 0.05, where stated). Confidence intervals for correlation and *R*^2^ comparisons were computed by patient-level bootstrap resampling with fixed random seeds.

#### Multiple-comparison correction

BH-FDR correction was applied in exactly two places: the Fig. 3 partial-correlation forest plot (corrected within each of 4 cohorts *×*2 targets, *q <* 0.05) and the Connection 1/Connection 2 analysis (corrected jointly across all 64 comparisons, *q <* 0.05). All other statistical comparisons in this manuscript are either single comparisons, small pre-specified nested-model comparisons interpreted via confidence-interval overlap, or explicitly uncorrected exploratory comparisons.

### Survival analysis

Survival analyses were performed in a baseline CVD-free dataset drawn from SHHS. Participants with prevalent myocardial infarction, stroke, congestive heart failure, or angina were excluded to evaluate prognostic associations in individuals without pre-existing cardiovascular disease. Participants with non-positive follow-up time were also excluded, yielding a final analytic cohort of 4,694 participants. Detailed cohort construction is provided in the Supplementary Information.

We evaluated three outcomes separately: all-cause mortality, CVD mortality, and non-CVD mortality. CVD mortality was defined using the cardiovascular- or coronary-heart-disease death classification provided by SHHS, and non-CVD mortality as all-cause death not classified as CVD/CHD death. For each outcome and each predictor (ΔAHI_brain_, AHI_brain,event-free_), Cox proportional-hazards models were fit with four levels of progressive covariate adjustment and with the predictor parameterized separately as quartiles and as a continuous per-SD term.

To compare their mortality associations with that of conventional AHI, crude and fully adjusted models were also fit separately for AHI_GT_, ΔAHI_brain_, and AHI_brain,event-free_ using the same clinical covariates. Discrimination was assessed using Harrell’s C-index, and the proportional-hazards assumption was evaluated using Schoenfeld residuals. Additional cohort-construction details and complete model-specific results are provided in Supplementary Note 3.

## Data availability

The KISS dataset cannot be made publicly available because it contains sensitive clinical polysomnography data subject to legal restrictions under the Personal Information Protection Act of the Republic of Korea. De-identified KISS data have been deposited with the Korea National Information Society Agency (NIA) AI Hub and may be requested through its controlled-access procedures for legitimate, non-commercial academic research, including replication of the findings reported here. Data-access inquiries should be sent to the corresponding authors, who will forward them for review under these procedures; access requires a data-use agreement and, where applicable, institutional review board approval, with decisions typically made within several weeks of a complete application.

MESA, MrOS, and SHHS data are available to qualified researchers through the National Sleep Research Resource (NSRR, https://sleepdata.org), subject to the data use agreements of the respective parent studies.

## Code availability

The code necessary to reproduce the findings of this study, including model training and evaluation scripts, is publicly available at https://github.com/hyunnie94/BrainAHI.

## Acknowledgements

The National Heart, Lung, and Blood Institute provided funding for the ancillary MrOS Sleep Study, “Outcomes of Sleep Disorders in Older Men,” under the following grant numbers: R01 HL071194, R01 HL070848, R01 HL070847, R01 HL070842, R01 HL070841, R01 HL070837, R01 HL070838, and R01 HL070839. The MESA Sleep ancillary study was funded by NIH-NHLBI Association of Sleep Disorders with Cardiovascular Health Across Ethnic Groups (RO1 HL098433); MESA itself is supported by NHLBI-funded contracts HHSN268201500003I, N01-HC-95159 through N01-HC-95169, and by cooperative agreements UL1-TR-000040, UL1-TR-001079, and UL1-TR-001420 funded by NCATS.The SHHS was supported by National Heart, Lung, and Blood Institute cooperative agreements U01HL53916 (University of California, Davis), U01HL53931 (New York University), U01HL53934 (University of Minnesota), U01HL53937 and U01HL64360 (Johns Hopkins University), U01HL53938 (University of Arizona), U01HL53940 (University of Washington), U01HL53941 (Boston University), and U01HL63463 (Case Western Reserve University). The National Sleep Research Resource was supported by the National Heart, Lung, and Blood Institute (R24 HL114473, 75N92019R002).

This work was supported by the AI-Bio Research Grant through Seoul National University; the Korea Health Technology R&D Project through the Korea Health Industry Development Institute (KHIDI), funded by the Ministry of Health & Welfare, Republic of Korea (grant number RS-2024-00439991); the SNUBH Research Fund (grant number 37-2025-2500); and the National AI Project of the Ministry of Science and ICT and the AIIS.

## Author contributions

H.K.L. contributed to the research design, performed the primary analyses and interpretation of results, and led the manuscript writing. K.P. and Y.R.C. contributed to model development and performed validation analyses, and both contributed to manuscript writing. D.L. contributed to the survival analysis. H.-S.K. co-supervised the project, assisted with study design, provided methodological and machine-learning supervision, and provided critical feedback on the manuscript. H.-W.S. conceived the overall study concept, supervised the clinical aspects of the work and provided access to the KISS cohort, and oversaw the entire project. H.-S.K. and H.-W.S. obtained funding. All authors reviewed and approved the final manuscript.

